# Nicotine pouch use for smoking cessation in Great Britain: real-world effectiveness, trends, and user characteristics, 2021–2026

**DOI:** 10.64898/2026.10.21.26366107

**Authors:** Sarah E. Jackson, Harry Tattan-Birch, Haoxiong Sun, Sharon Cox, Melissa Oldham, Jamie Brown

## Abstract

**Objective:** To examine trends in use of nicotine pouches as a smoking cessation aid, characteristics of people using pouches during smoking quit attempts, the characteristics of quit attempts involving pouches, and the real-world association between pouch use and quit success.

**Design:** Monthly cross-sectional survey.

**Setting:** Great Britain.

**Participants:** 7,995 adults (≥18 years) who reported making a serious attempt to stop smoking in the past year, surveyed between June 2021 and July 2026.

**Main outcome measures:** Participants reported use of nicotine pouches during the most recent smoking quit attempt (yes/no). Quit success was defined as reporting that the participant was still not smoking following the attempt. Associations between pouch use and quit success were estimated using logistic regression, adjusting for sociodemographic characteristics, use of other cessation aids, characteristics of the quit attempt, strength of urges to smoke, and survey year.

**Results:** Among adults who attempted to quit smoking, the proportion using pouches as a cessation aid increased from 2.4% [95%CI 1.3-4.1%] in June 2021 to 6.7% [4.6-9.6%] in July 2026 (prevalence ratio [PR]=2.83 [1.37-5.81]), with particularly large increases among 18-24-year-olds (2.2% to 18.9%; PR=8.75 [4.15-26.79]) and men (1.8% to 11.0%; PR=6.05 [3.19-14.31]). In 2024-26, 43.8% of those using pouches as a cessation aid were aged 18-24 and 69.5% were men. Pouch users also commonly reported using e-cigarettes (49.6%) or over-the-counter nicotine replacement therapy (37.9%) during the same quit attempt. Unadjusted quit success was 33.5% [27.1-39.9] among pouch users compared with 27.0% [25.9-28.1] among those who did not use pouches (OR=1.36 [1.02-1.82]). After adjustment, the association was attenuated (OR_adj_=1.15 [0.80-1.65]), with the 95%CI compatible with both a modest adverse association and a modest to large benefit. The association differed according to concurrent e-cigarette use (*p*_interaction_=0.002): among those not using e-cigarettes as a cessation aid, pouch use was associated with higher quit success (OR_adj_=1.91 [1.23-2.97]), whereas evidence of an association among e-cigarette users was uncertain (OR_adj_=0.63 [0.36-1.11]).

**Conclusions:** Use of nicotine pouches as a smoking cessation aid increased substantially in Great Britain between 2021 and 2026, particularly among younger adults and men. Pouches were frequently used alongside other cessation aids. While the overall association with quit success was uncertain, pouch use was associated with higher quit success among those not using e-cigarettes. If causal, this suggests a potential role for pouches among people not already using an established cessation aid.

## Introduction

Use of oral nicotine pouches has increased rapidly in Great Britain since 2020, particularly among younger adults, and especially young men.^1,2^ Nicotine pouches are small, tobacco-free products containing nicotine that are placed between the gum and lip, where nicotine is absorbed through the oral mucosa.^3^ Several major tobacco companies now manufacture and market nicotine pouches, which have become a prominent part of the expanding nicotine market.^4,5^

Recent evidence suggests that pouches are increasingly being used in the context of smoking cessation, with the proportion of smoking quit attempts involving pouches rising from around 2% between 2021 and 2022 to 6.5% by March 2025.^1^ Although this remains lower than the use of e-cigarettes (∼35% in April 2025) or over-the-counter nicotine replacement therapy (NRT; ∼16%),^6^ pouches are now used in a proportion of smoking quit attempts comparable to, or higher than, several prescription pharmacotherapies and behavioural support interventions delivered through stop smoking services.^7^

This increase raises important questions about their role for smoking cessation. Nicotine pouches are not licensed as smoking cessation medicines and were largely unregulated during the study period. They are now being brought under consumer regulation through the Tobacco and Vapes Act 2026,^8^ so evidence on whether pouches support smoking cessation, and who is using them for this purpose, is directly relevant to how these regulations are designed.

It is unclear whether pouches’ growing use reflects substitution for other cessation aids or complementary use alongside them. Evidence on their effectiveness for smoking cessation is currently limited. Randomised controlled trial evidence is lacking,^9^ and observational evidence remains sparse. In a previous analysis using Smoking Toolkit Study data up to mid-2024,^7^ people who used nicotine pouches during a smoking quit attempt had a relatively high unadjusted quit rate (30.1%; 95% CI 19.3–40.8%). However, because only a small number of participants reported using pouches in quit attempts (*n*=84), the adjusted association with cessation was imprecisely estimated (OR=1.21, 95% CI 0.70–2.07). Although the point estimate suggested a modest benefit, the wide confidence interval was compatible with effects ranging from a modest reduction to a benefit as large as those observed for some of the most effective established cessation aids, including varenicline (OR=1.80) and e-cigarettes (OR=1.95).^7^ Substantial growth in pouch use since then^1^ provides an opportunity to obtain a more precise estimate of their real-world effectiveness and to provide a more detailed picture of how pouches are being used in practice.

Little is also known about who uses nicotine pouches when attempting to quit smoking. It is unclear whether patterns of use mirror those observed for nicotine pouch use more generally, such as higher uptake among younger adults and men^1^, or differ in the context of cessation. There is also limited evidence on how people who use pouches differ from other people attempting to stop smoking, including in their sociodemographic characteristics and features of the quit attempt, such as whether quitting was planned or abrupt and whether other cessation aids are used concurrently.

This study aimed to provide a comprehensive assessment of nicotine pouch use for smoking cessation in Great Britain between 2021 and 2026. Specifically, we aimed to:

1. Estimate the real-world effectiveness of nicotine pouches for smoking cessation using an additional two years of data to improve the precision and robustness of our previous estimate.7
2. Examine the extent to which nicotine pouch use in quit attempts, and trends over time, vary across key sociodemographic subgroups (age, gender, and socioeconomic position).
3. Describe the profile of those making a quit attempt who used nicotine pouches in 2024-26, compared with (a) the overall population of adults making a quit attempt over the same period, (b) those who did not use nicotine pouches, and (c) those who used other cessation aids (e-cigarettes, over-the-counter NRT, prescription pharmacotherapy, or face-to-face behavioural support), including their sociodemographic characteristics, strength of urges to smoke, features of the most recent quit attempt, and concurrent use of other cessation aids.

## Methods

### Pre-registration

The study protocol and analysis plan were pre-registered on Open Science Framework (osf.io/a8gvq).

### Design

Data were drawn from the Smoking Toolkit Study, an ongoing monthly cross-sectional survey of a representative sample of adults (≥16 years) in Great Britain.^10,11^ The study uses a hybrid of random probability and simple quota sampling to select a new sample of approximately 2,450 adults each month. Data are collected through telephone interviews. Raking is used to weight the sample to match the population.^10^ This profile is determined each month by combining data from the UK Census, the Office for National Statistics mid-year estimates, and the annual National Readership Survey.^10^ Comparisons with other national household surveys and sales data indicate the survey achieves nationally representative estimates of key sociodemographic and nicotine use variables.^10,12^

### Power calculation for real-world effectiveness analysis

Based on a quit rate of ∼20% overall in the Smoking Toolkit Study and an average of ∼3.5% of participants using pouches in quit attempts over the whole period, we calculated that we would require a sample of at least 7,575 past-year quit attempts in order to have 80% power to detect an OR of 1.5. We considered an OR of 1.5 to be a plausible and clinically meaningful effect size in the absence of randomised evidence, and is consistent with the range of effect sizes that remained compatible with the confidence interval from our previous observational analysis.^7^

### Sample

We analysed data collected monthly between June 2021 (the first wave to assess use of nicotine pouches in smoking quit attempts) and July 2026. We included all participants aged ≥18 years who reported having attempted to quit smoking in the past 12 months, providing a sample of 7,995 participants.

### Measures

Full details of the measures are provided in the study protocol (osf.io/a8gvq).

Cessation aids used were assessed by asking participants which aids they used during their most recent serious attempt to quit smoking. Use of nicotine pouches was categorised as yes/no. We also analysed use of the other aids, including e-cigarettes, over-the-counter nicotine replacement therapy (NRT), prescription pharmacotherapy (NRT and non-nicotine medications varenicline, bupropion, and cytisine), and face-to-face behavioural support; these aids were selected because they are commonly used and/or have demonstrated effectiveness for increasing quit success rates.^7,13–16^

Other variables which are known to influence quit success were assessed, including time since the attempt began, number of prior smoking quit attempts in the past year, whether the attempt was planned, and whether participants cut down first or quit abruptly. Those who responded ‘don’t know’ to any of these questions were coded as missing in the primary analysis and included as a separate category in a sensitivity analysis.

Strength of urges to smoke over the last 24 hours was assessed using a validated two-item measure.^17^ Participants reporting any urges rated their strength from slight to extremely strong, while those reporting no urges were assigned a score of 0. Scores ranged from 0 to 5, with higher scores indicating stronger urges. This measure has comparable predictive validity for cessation to the Fagerström Test of Cigarette Dependence and Heaviness of Smoking Index.^18^

Quit success was defined as reporting that they were ‘still not smoking’ following their most recent serious quit attempt. As participants were surveyed at varying intervals after the attempt, this measure reflects abstinence of varying duration rather than abstinence sustained for a fixed period.

Sociodemographic characteristics assessed included age, gender, and occupational social grade. Age was categorised into six groups for descriptive analyses and modelled continuously using restricted cubic splines when estimating age-specific prevalence and trends in nicotine pouch use to allow for non-linear associations. Gender was categorised as man, woman, or other, with the latter group excluded from gender-stratified trend analyses due to small numbers but included in other analyses. Social grade was measured using the National Readership Survey classification^19^ and categorised as ABC1 (includes managerial, professional, and upper supervisory occupations) or C2DE (includes manual routine, semi-routine, lower supervisory, state pension, and long-term unemployed).

Time was assessed using survey wave, coded continuously (June 2021 = 1 through July 2026 = 62) and modelled non-linearly using splines (see *Statistical analysis*). Survey year was categorised from 2021 to 2026.

### Statistical analysis

Analyses were conducted in R v.4.6.0 using weighted data. Missing cases were excluded on a per-analysis basis.

### Real-world effectiveness

Using data pooled across the period (June 2021 to July 2026), we estimated the association between nicotine pouch use during the most recent smoking quit attempt and quit success. First, we calculated the unadjusted quit success rate (with 95% CI) among participants who attempted to quit with and without the use of nicotine pouches. We then fitted unadjusted and adjusted multivariable logistic regression models with quit success (abstinent vs. not) as the outcome and nicotine pouch use (yes/no) as the primary exposure. The adjusted model included sociodemographic characteristics (age, gender, social grade), use of other cessation aids (e-cigarettes, over-the-counter NRT, prescription pharmacotherapy, face-to-face behavioural support), other features of the smoking quit attempt (time since the quit attempt started, number of past-year smoking quit attempts, planned/unplanned, abrupt/gradual), strength of urges to smoke, and survey year.

We conducted three unplanned sensitivity analyses. First, we repeated the adjusted model retaining participants who reported that they did not know when their quit attempt started or whether it was planned or unplanned (coding don’t know responses as an additional category), rather than excluding them because of missing information on these variables. Second, we repeated the analysis excluding strength of urges to smoke from the covariates, because this measure may mediate the association between nicotine pouch use and quit success and was assessed at the time of the survey rather than at the start of the quit attempt. Third, because concurrent e-cigarette use was common among participants who used nicotine pouches as a cessation aid, we examined whether pouches were associated with quit success differently among those who did and did not use e-cigarettes, given that any additional benefit of pouches might be limited among those already using an established cessation aid.^15^ We added a nicotine pouch use-by-e-cigarette use interaction term to the adjusted model and estimated associations separately among participants who did and did not report e-cigarette use during their quit attempt.

### Differences in use by sociodemographic characteristics

Using data pooled across the period (June 2021 to July 2026), we examined differences in the prevalence of nicotine pouch use in smoking quit attempts according to age, gender, and social grade. To estimate overall differences between sociodemographic groups, we fitted logistic regression models with nicotine pouch use (yes/no) as the outcome, including each sociodemographic characteristic as the exposure of interest. Results are reported as percentages and prevalence ratios (PRs) with 95% CIs, with PRs and CIs calculated using bootstrapping with 1,000 replications.

### Time trends

To assess whether trends over time differed across sociodemographic groups, we fitted separate logistic regression models including an interaction between time (survey wave) and each sociodemographic characteristic (age, gender, and social grade). We also modelled the overall trend among all adults who made a quit attempt using a model only including time as a predictor, updating the trend we reported previously^1^ with an additional 16 months of data. Time was modelled as a continuous variable using restricted cubic splines to allow for non-linear trends. The number of knots (three to four) for each model was selected by comparing Akaike Information Criterion (AIC) values, with preference given to the simplest model within two AIC units of the model with the lowest AIC (see **Table S1** for details of model selection).^20^ Age was also modelled using splines with three knots at the 0%, 50%, and 95% quantiles.

Predicted probabilities of nicotine pouch use were estimated from the fitted models and plotted over time for all adults (time-only model) and for each sociodemographic subgroup (interaction models). Absolute changes in prevalence over the study period were calculated as the difference between the predicted prevalence at the start and end of the study period, and PRs were calculated as the ratio of the predicted prevalence at the end of the study period to that at the start. 95% CIs for these change estimates were calculated using bootstrapping with 1,000 replications.

Because the age- and gender-specific models suggested substantial differences in trends, we conducted an unplanned exploratory analysis in which we fitted a further logistic regression model including a three-way interaction between time, age, and gender. We used predicted estimates from this model to plot trends in each age group separately for men and women.

### Profile comparisons

Using data from January 2024 onwards, we described the characteristics of: 1) all adults who made a smoking quit attempt; 2) those who used nicotine pouches during their most recent quit attempt; 3) those who did not use nicotine pouches during their most recent quit attempt; and 4) those who did not use nicotine pouches but used other cessation aids (e-cigarettes, over-the-counter NRT, prescription pharmacotherapy, face-to-face behavioural support). We restricted this analysis to recent data to provide an up-to-date description of nicotine pouch users while reducing confounding by time (given the sharp rise in pouch use in quit attempts,^1^ pouch user data was heavily skewed to recent years). Profiles were summarised using descriptive statistics (percentages and 95% CIs) across key sociodemographic characteristics (age, gender, social grade), features of the smoking quit attempt (abrupt/gradual, planned/unplanned), use of cessation aids (nicotine pouches, e-cigarettes, over-the-counter NRT, prescription pharmacotherapy, face-to-face behavioural support), and strength of urges to smoke.

## Results

A total of 142,299 adults aged ≥18 years in Great Britain were surveyed between June 2021 and July 2026. Of 23,406 (16.4%) participants who reported smoking in the past year, 7,995 (34.2%) reported having made at least one serious attempt to stop smoking over this period and formed our analytic sample. Sample characteristics and missing data on each variable are summarised in **Tables S2-S3**.

### Real-world effectiveness

Among all adults who reported a past-year smoking quit attempt across the study period (*n*=7,995), 3.9% [95% CI 3.4–4.4] reported using nicotine pouches as a cessation aid. The unadjusted quit success rate was 33.5% [27.1–39.9] among those who used pouches during their quit attempt and 27.0% [25.9–28.1] among those who did not, corresponding to an unadjusted OR of 1.36 [95% CI 1.02–1.82].

After adjustment for sociodemographic characteristics, use of other cessation aids, features of the quit attempt, strength of urges to smoke, and survey year, the association was attenuated and there was no clear evidence of an association between nicotine pouch use and quit success. Among participants with complete data on all covariates (*n*=7,325), the adjusted OR was 1.15 [95% CI 0.80–1.65]. Results were similar in a sensitivity analysis that retained participants who reported that they did not know when their quit attempt started or whether it was planned or abrupt (*n*=7,816; adjusted OR=1.11, 95% CI 0.78–1.58). When strength of urges to smoke was excluded from the model, the association was somewhat larger but remained imprecisely estimated (*n*=7,428; adjusted OR=1.28, 95% CI 0.92–1.77).

Because concurrent e-cigarette use was common among pouch users (see *Profile comparisons* below), we also examined whether the association differed according to e-cigarette use. There was evidence of an interaction between use of pouches and e-cigarettes as smoking cessation aids on quit success (*n*=7,325; interaction OR=0.32, 95% CI 0.16–0.65, *p*=0.002), with a stronger positive association among participants who did not use e-cigarettes (*n*=4,749; adjusted OR=1.91, 95% CI 1.23–2.97) whereas, among those using e-cigarettes, the estimated association was in the opposite direction, although the confidence interval was compatible with both lower and slightly higher odds of quit success (*n*=2,576; adjusted OR=0.63, 95% CI 0.36–1.11). Unadjusted quit success showed a similar pattern: among participants who did not use e-cigarettes as a cessation aid, quit success was 38.4% [29.5–47.3] among pouch users (*n*=154) compared with 24.0% [22.7–25.4] among non-users (*n*=5,058); among those who used e-cigarettes, corresponding estimates were 27.8% [18.8–36.8] among pouch users (*n*=126) and 32.2% [30.2–34.2] among non-users (*n*=2,657).

### Differences in use by sociodemographic characteristics

Across the study period, nicotine pouch use varied substantially by age and gender but showed little evidence of socioeconomic differences (**Table 1**). Prevalence was highest among those aged 18–24 years and declined progressively with age (PR vs ≥65 years 5.00, 95% CI 2.80–8.89). Men were approximately twice as likely as women to report using pouches in smoking quit attempts (PR 2.05, 95% CI 1.51–2.67). Prevalence was also relatively high among adults reporting another gender (8.8% [3.6–14.0]; PR vs. women 3.69 [1.67–6.14]), although estimates were imprecise. There was little evidence of an overall difference by occupational social grade (PR 1.14, 95% CI 0.88–1.46).

**Table 1.** Prevalence of nicotine pouch use as a smoking cessation aid among adults in Great Britain who made a past-year smoking quit attempt, 2021-26 (aggregated across monthly survey waves)

|  | % [95% CI] | PR [95% CI]<sup>1</sup> |
| --- | --- | --- |
| Overall | 3.9 [3.4–4.4] | - |
| Age (years)<sup>2</sup> |  |  |
| 18-24 | 7.5 [6.2–8.8] | 5.00 [2.80–8.89] |
| 25-34 | 3.9 [3.2–4.6] | 2.61 [1.32–4.94] |
| 35-44 | 2.4 [1.9–2.9] | 1.58 [0.78–2.93] |
| 45-54 | 1.9 [1.5–2.3] | 1.21 [0.67–2.06] |
| 55-64 | 1.7 [1.3–2.2] | 1.07 [0.73–1.55] |
| ≥65 | 1.7 [0.8–2.6] | Ref |
| Gender |  |  |
| Woman | 2.5 [1.9–3.1] | Ref |
| Man | 5.0 [4.2–5.8] | 2.05 [1.51–2.67] |
| Other | 8.8 [3.6–14.0] | 3.69 [1.67–6.14] |
| Social grade |  |  |
| ABC1 (more advantaged) | 4.1 [3.5–4.8] | 1.14 [0.88–1.46] |
| C2DE (less advantaged) | 3.7 [2.9–4.4] | Ref |

CI, confidence interval. PR, prevalence ratio. Ref, reference category.
<sup>1</sup> Estimated from logistic regression models adjusted for survey year, using bootstrapping.
<sup>2</sup> Age modelled continuously using restricted cubic splines using data from people of all ages. Estimates for each age group were taken from the mean exact age of individuals in that age range.

### Time trends

Among those making a past-year quit attempt, nicotine pouch use as a smoking cessation aid increased substantially over the study period, from a modelled 2.4% [95% CI 1.3–4.1] in June 2021 to 6.7% [4.6–9.6] in July 2026 (PR=2.83, 95% CI 1.37–5.81; **Table 2**). This change was non-linear: prevalence remained relatively stable at around 2.1% up to April 2023, before rising steadily to 5.8% by April 2025. The rate of increase subsequently slowed, with prevalence reaching 6.7% by July 2026 (**Figure 1A**).

**Figure 1.**
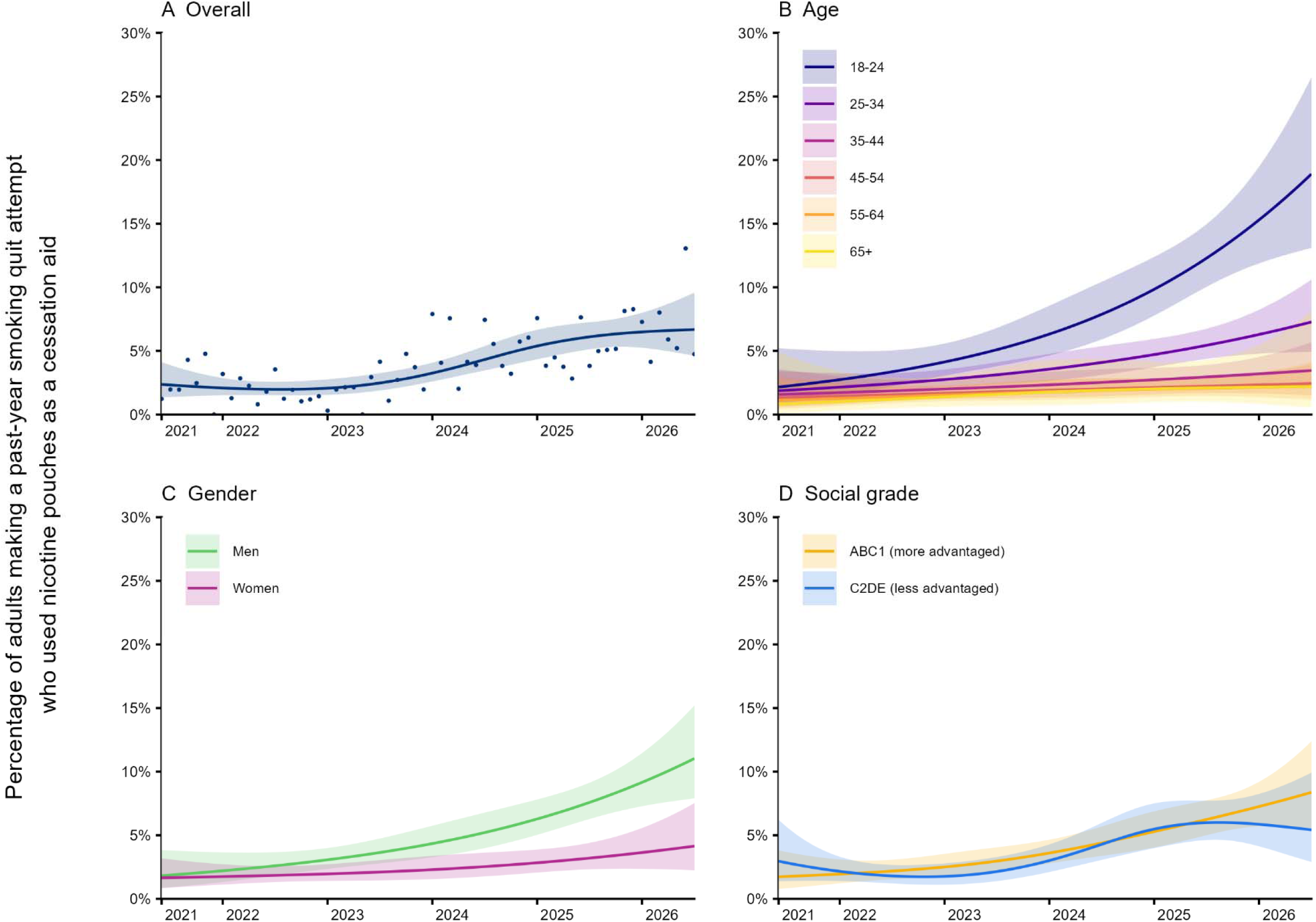
Percentage of adults making a past-year smoking quit attempt who used nicotine pouches as a cessation aid, overall and across subgroups in Great Britain, 2021-2026. Lines represent the modelled weighted prevalence by monthly survey wave (estimated using logistic regression, with time modelled with restricted cubic splines; see Table S1 for model selection), shaded bands represent 95% CIs, and points represent the unmodelled weighted prevalence in each month. See **Table 2** for exact estimates in the first and last months of this period.

**Table 2.**
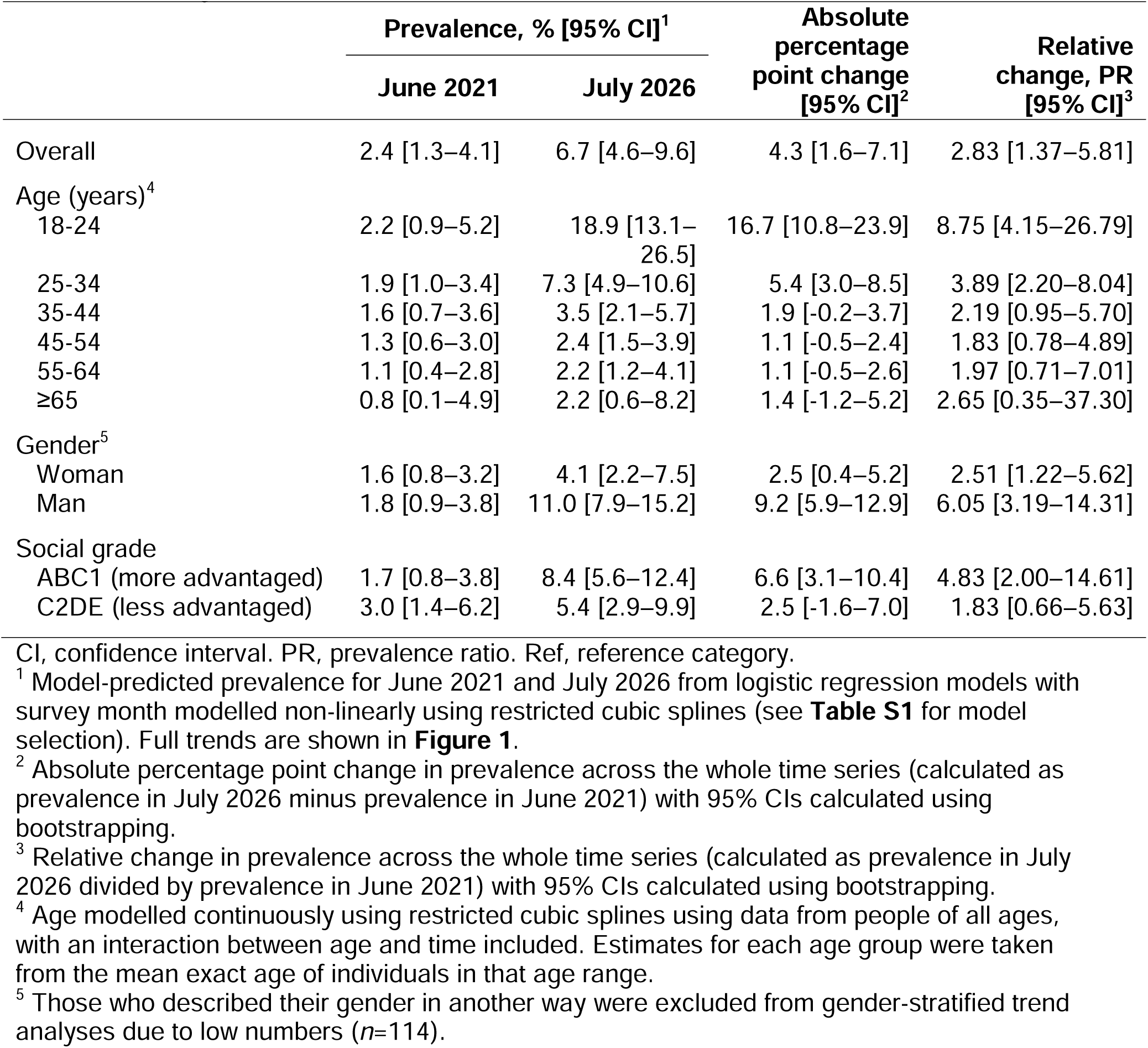
Modelled estimates of the change in prevalence of nicotine pouch use as a smoking cessation aid among adults in Great Britain who made a past-year smoking quit attempt from June 2021 to July 2026

|  | Prevalence, % [95% CI]<sup>1</sup> |  | Absolute percentage point change [95% CI]<sup>2</sup> | Relative change, PR [95% CI]<sup>3</sup> |
| --- | --- | --- | --- | --- |
|  | June 2021 | July 2026 |  |  |
| Overall | 2.4 [1.3–4.1] | 6.7 [4.6–9.6] | 4.3 [1.6–7.1] | 2.83 [1.37–5.81] |
| Age (years)<sup>4</sup> |  |  |  |  |
| 18-24 | 2.2 [0.9–5.2] | 18.9 [13.1–26.5] | 16.7 [10.8–23.9] | 8.75 [4.15–26.79] |
| 25-34 | 1.9 [1.0–3.4] | 7.3 [4.9–10.6] | 5.4 [3.0–8.5] | 3.89 [2.20–8.04] |
| 35-44 | 1.6 [0.7–3.6] | 3.5 [2.1–5.7] | 1.9 [-0.2–3.7] | 2.19 [0.95–5.70] |
| 45-54 | 1.3 [0.6–3.0] | 2.4 [1.5–3.9] | 1.1 [-0.5–2.4] | 1.83 [0.78–4.89] |
| 55-64 | 1.1 [0.4–2.8] | 2.2 [1.2–4.1] | 1.1 [-0.5–2.6] | 1.97 [0.71–7.01] |
| ≥65 | 0.8 [0.1–4.9] | 2.2 [0.6–8.2] | 1.4 [-1.2–5.2] | 2.65 [0.35–37.30] |
| Gender<sup>5</sup> |  |  |  |  |
| Woman | 1.6 [0.8–3.2] | 4.1 [2.2–7.5] | 2.5 [0.4–5.2] | 2.51 [1.22–5.62] |
| Man | 1.8 [0.9–3.8] | 11.0 [7.9–15.2] | 9.2 [5.9–12.9] | 6.05 [3.19–14.31] |
| Social grade |  |  |  |  |
| ABC1 (more advantaged) | 1.7 [0.8–3.8] | 8.4 [5.6–12.4] | 6.6 [3.1–10.4] | 4.83 [2.00–14.61] |
| C2DE (less advantaged) | 3.0 [1.4–6.2] | 5.4 [2.9–9.9] | 2.5 [-1.6–7.0] | 1.83 [0.66–5.63] |

CI, confidence interval. PR, prevalence ratio. Ref, reference category.
<sup>1</sup> Model-predicted prevalence for June 2021 and July 2026 from logistic regression models with survey month modelled non-linearly using restricted cubic splines (see **Table S1** for model selection). Full trends are shown in **Figure 1**.
<sup>2</sup> Absolute percentage point change in prevalence across the whole time series (calculated as prevalence in July 2026 minus prevalence in June 2021) with 95% CIs calculated using bootstrapping.
<sup>3</sup> Relative change in prevalence across the whole time series (calculated as prevalence in July 2026 divided by prevalence in June 2021) with 95% CIs calculated using bootstrapping.
<sup>4</sup> Age modelled continuously using restricted cubic splines using data from people of all ages, with an interaction between age and time included. Estimates for each age group were taken from the mean exact age of individuals in that age range.
<sup>5</sup> Those who described their gender in another way were excluded from gender-stratified trend analyses due to low numbers ( $n=114$ ).

The increase was particularly pronounced among younger adults (**Figure 1B**). Prevalence increased from 2.2% to 18.9% among those aged 18–24 years (PR=8.75, 95% CI 4.15–26.79) and from 1.9% to 7.3% among those aged 25–34 years (PR=3.89, 95% CI 2.20–8.04; **Table 2**). There was less evidence of an increase among adults aged ≥35 years: although point estimates suggested approximately two-fold relative increases across these age groups (PR range 1.83–2.65), 95% CIs were wide and included the possibility of no change. Absolute prevalence remained low in these groups, increasing from around 1–2% at the start of the study period to around 2–4% at the end.

The increase was also greater among men than women, although prevalence increased in both groups (**Figure 1C**). Among men, prevalence increased from 1.8% to 11.0% (PR=6.05, 95% CI 3.19–14.31), compared with an increase from 1.6% to 4.1% among women (PR=2.51, 95% CI 1.22–5.62; **Table 2**). The largest absolute increase was observed among 18–24-year-old men, among whom prevalence increased from 3.1% to 26.1% (**Figure S1; Table S4**).

Patterns also differed by occupational social grade (**Figure 1D**). Prevalence increased substantially among adults in more advantaged social grades, from 1.7% to 8.4% (PR=4.83, 95% CI 2.00–14.61), whereas the increase was smaller and more uncertain among those in less advantaged grades (3.0% to 5.4%; PR=1.83, 95% CI 0.66–5.63; **Table 2**). Prevalence initially increased in both groups at a roughly similar rate, but subsequently levelled off among less advantaged adults while continuing to increase among those who were more advantaged (**Figure 1D**).

### Profile comparisons

Among adults who made a smoking quit attempt between 2024 and 2026 (*n*=3,898), 5.6% [4.7–6.4] reported using nicotine pouches as a cessation aid. This was higher than the proportion using non-nicotine prescription pharmacotherapy (varenicline, bupropion, or cytisine; 2.1%), similar to the proportion using face-to-face behavioural support (5.5%), and lower than the proportions using prescription NRT (7.2%), over-the-counter NRT (17.9%), or e-cigarettes (37.9%; **Table 3**). **Table 3** also compares the sociodemographic characteristics, features of the quit attempt, concurrent use of other cessation aids, and strength of urges to smoke among those who did and did not use pouches.

**Table 3.** Profile of adults who attempted to quit smoking, 2024-26.

|  | Column % [95% CI]<sup>1</sup> |  |  |  |
| --- | --- | --- | --- | --- |
|  | All adults who made a smoking quit attempt | Used nicotine pouches | Did not use nicotine pouches | Did not use nicotine pouches but used other cessation aids<sup>2</sup> |
| Unweighted *n* | 3,898 | 196 | 3,702 | 1,976 |
| Age (years) |  |  |  |  |
| Mean (SD) | 39.4 (16.0) | 30.1 (13.1) | 40.0 (15.9) | 40.7 (15.6) |
| 18-24 | 22.2 [20.7–23.8] | 43.8 [36.1–51.8] | 21.0 [19.4–22.6] | 19.6 [17.5–21.8] |
| 25-34 | 26.6 [25.0–28.3] | 35.0 [27.6–43.1] | 26.1 [24.5–27.8] | 24.6 [22.4–26.9] |
| 35-44 | 17.3 [15.9–18.7] | 8.3 [5.0–13.4] | 17.8 [16.4–19.3] | 17.7 [15.8–19.7] |
| 45-54 | 13.0 [11.9–14.2] | 4.7 [2.2–9.6] | 13.5 [12.3–14.8] | 15.5 [13.8–17.4] |
| 55-64 | 12.0 [11.0–13.2] | 4.9 [2.8–8.6] | 12.5 [11.4–13.6] | 14.4 [12.8–16.1] |
| ≥65 | 8.8 [8.0–9.7] | 3.4 [1.7–6.6] | 9.2 [8.3–10.1] | 8.3 [7.2–9.5] |
| Gender |  |  |  |  |
| Man | 52.3 [50.5–54.1] | 69.5 [61.6–76.4] | 51.3 [49.4–53.1] | 46.1 [43.6–48.6] |
| Woman | 46.5 [44.7–48.3] | 27.4 [20.7–35.3] | 47.6 [45.8–49.5] | 52.8 [50.3–55.3] |
| Other | 1.2 [0.9–1.6] | 3.0 [1.5–6.1] | 1.1 [0.8–1.5] | 1.1 [0.7–1.6] |
| Social grade |  |  |  |  |
| ABC1 (more advantaged) | 41.9 [40.2–43.6] | 44.3 [36.8–52.0] | 41.8 [40.0–43.5] | 39.2 [36.9–41.5] |
| C2DE (less advantaged) | 58.1 [56.4–59.8] | 55.7 [48.0–63.2] | 58.2 [56.5–60.0] | 60.8 [58.5–63.1] |
| Quit attempt was... |  |  |  |  |
| Abrupt | 51.4 [49.6–53.2] | 50.6 [42.6–58.6] | 51.5 [49.6–53.3] | 46.6 [44.0–49.1] |
| Gradual | 48.6 [46.8–50.4] | 49.4 [41.4–57.4] | 48.5 [46.7–50.4] | 53.4 [50.9–56.0] |
| Planned | 40.3 [38.5–42.1] | 35.5 [28.3–43.4] | 40.6 [38.7–42.4] | 45.8 [43.3–48.4] |
| Unplanned | 59.7 [57.9–61.5] | 64.5 [56.6–71.7] | 59.4 [57.6–61.3] | 54.2 [51.6–56.7] |
| Quit attempt involved the use of... |  |  |  |  |
| Nicotine pouches | 5.6 [4.8–6.5] | 100 | 0 | 0 |
| E-cigarettes | 37.9 [36.1–39.6] | 49.6 [41.7–57.5] | 37.2 [35.4–39.0] | 69.3 [66.9–71.5] |
| NRT bought over-the-counter | 17.9 [16.6–19.3] | 37.9 [30.5–46.0] | 16.7 [15.4–18.2] | 31.2 [28.9–33.6] |
| Prescription pharmacotherapy<sup>3</sup> | 8.7 [7.7–9.7] | 8.6 [5.1–14.2] | 8.7 [7.7–9.7] | 16.2 [14.4–18.1] |
| NRT | 7.2 [6.4–8.2] | 8.0 [4.6–13.6] | 7.2 [6.3–8.2] | 13.4 [11.9–15.1] |
| Non-nicotine pharmacotherapy<sup>4</sup> | 2.1 [1.6–2.7] | 2.6 [1.0–6.6] | 2.1 [1.6–2.7] | 3.9 [3.0–5.1] |
| Face-to-face behavioural support | 5.5 [4.8–6.4] | 6.9 [3.9–12.0] | 5.5 [4.7–6.3] | 10.2 [8.8–11.7] |
| Strength of urges to smoke |  |  |  |  |
| Mean (SD) | 1.5 (1.3) | 1.4 (1.4) | 1.5 (1.3) | 1.6 (1.4) |
| 0. Not at all | 29.7 [28.0–31.4] | 38.7 [31.1–46.8] | 29.2 [27.5–30.9] | 26.9 [24.7–29.3] |
| 1. Slight | 19.3 [17.9–20.8] | 16.9 [11.7–23.6] | 19.5 [18.0–21.0] | 19.1 [17.1–21.2] |
| 2. Moderate | 29.0 [27.4–30.7] | 25.6 [19.4–33.0] | 29.2 [27.5–30.9] | 29.5 [27.3–31.9] |
| 3. Strong | 14.4 [13.2–15.7] | 11.8 [7.5–18.2] | 14.6 [13.3–15.9] | 15.6 [13.8–17.5] |
| 4. Very strong | 4.7 [4.0–5.5] | 3.1 [1.5–6.4] | 4.8 [4.1–5.7] | 5.4 [4.4–6.6] |
| 5. Extremely strong | 2.9 [2.3–3.5] | 3.9 [1.8–8.4] | 2.8 [2.3–3.5] | 3.5 [2.7–4.6] |

<sup>1</sup> Unless otherwise specified. <sup>2</sup> E-cigarettes, over-the-counter NRT, prescription pharmacotherapy, or face-to-face behavioural support, but not nicotine pouches. <sup>3</sup> NRT, varenicline, bupropion, or cytisine. <sup>4</sup> Varenicline, bupropion, or cytisine.

Compared with those who did not use nicotine pouches in their quit attempt, those who used pouches were on average 10 years younger and a higher proportion were men (69.5% vs. 51.3%), but the distribution by social grade was similar. A higher proportion of pouch users reported no urges to smoke in the past 24 hours (38.7% vs. 29.2%). Use of other cessation aids was also common among pouch users: 49.6% reported using e-cigarettes and 37.9% over-the-counter NRT, compared with 37.2% and 16.7%, respectively, among those who did not use pouches.

Comparisons with those who did not use pouches but used other cessation aids showed a similar pattern. Pouch users were younger and a higher proportion were men, quit successfully, and reported no urges to smoke. In addition, a higher proportion reported that their quit attempt was unplanned (64.5% vs. 54.2%).

## Discussion

Among adults in Great Britain attempting to quit smoking, use of nicotine pouches as a cessation aid increased substantially between 2021 and 2026, particularly among younger adults and men. By July 2026, around one in every 15 quit attempts involved nicotine pouches, often alongside other cessation aids. Despite this growing use, it remains uncertain whether pouches help people quit smoking. Although quit success was higher among pouch users in unadjusted analyses, this difference was attenuated after adjusting for sociodemographic characteristics, use of other cessation aids, features of the smoking quit attempt, strength of urges to smoke, and survey year, meaning the independent association was uncertain. However, pouch use was associated with higher quit success among those not using e-cigarettes.

Our findings on the growth and demographic patterning of pouch use are broadly consistent with population-level evidence from Great Britain. We have previously shown that overall pouch use has increased rapidly since 2020, particularly among younger people and men, and that pouch use often occurs alongside smoking and vaping.^1,2^ We found a similar pattern among people attempting to quit smoking: by July 2026, almost one in five 18–24-year-olds making a quit attempt, and more than one in ten men, reported using nicotine pouches as a cessation aid. Around half of pouch users also used e-cigarettes during the same quit attempt, and over a third used over-the-counter NRT. There was little evidence of a difference in pouch use by social grade, although use for this purpose appeared to be recently increasing more rapidly among more advantaged adults who smoke and levelling off among those who are less advantaged. Whether socioeconomic differences in pouch uptake are emerging therefore warrants monitoring, particularly given existing inequalities in smoking and cessation.^21–23^

Our findings also provide insight into how nicotine pouches are used in real-world smoking quit attempts. Pouch users were somewhat more likely to report that their quit attempt was unplanned than those who used other cessation aids without pouches. This could reflect a pattern whereby people start using pouches while still smoking and then decide to quit, rather than starting use of pouches as part of a planned quit attempt, as has previously been observed among people who smoke and vape.^24^ However, our data cannot establish whether pouch use came before or after the decision to quit. Pouches were also often used alongside other nicotine products, suggesting that they are commonly part of a broader approach to quitting rather than used as a standalone intervention. Pouches were used in a similar proportion of quit attempts as face-to-face behavioural support and more commonly than effective non-nicotine prescription pharmacotherapies such as varenicline, bupropion, and cytisine,^14,16^ indicating that they have already become a relatively established component of the cessation landscape despite the limited evidence on their effectiveness. Restrictions on flavours, nicotine strength, and packaging that may be implemented under the Tobacco and Vapes Act 2026 could affect the appeal of pouches to adults attempting to quit, and continued monitoring will be needed if and when these measures take effect.

The concentration of pouch use among younger adults is important when interpreting the findings on effectiveness. In 2024-26, 79% of adults who used nicotine pouches as a smoking cessation aid were aged under 35, with 44% aged 18–24 years. Younger people who smoke differ in important ways from those who are older, including having a much higher prevalence of non-daily smoking.^25,26^ Pouch users may therefore differ from other people attempting to quit in their underlying likelihood of success, regardless of the cessation aid they use. Younger, less dependent smokers may find it easier to quit once they make an attempt, which could partly explain the higher unadjusted quit success among pouch users. The relatively low uptake of pouches among middle-aged and older adults also means that there is currently limited evidence about their effectiveness in these groups. As use becomes more widespread, it will be important to assess effectiveness among groups with higher levels of smoking dependence.

The concurrent use of other cessation aids is another important consideration. Nicotine pouches were often used alongside e-cigarettes and over-the-counter NRT rather than on their own. Although we adjusted for use of other cessation aids, we did not measure how these products were used within the same quit attempt, for example, how often, for how long, or in what order. Therefore, how much any association with cessation was due specifically to nicotine pouches rather than their use alongside other aids is unknown. People who are particularly motivated to quit or who struggle to maintain abstinence may be more likely to use multiple cessation aids, which could contribute to differences in quit success between pouch users and non-users. The high prevalence of concurrent e-cigarette use among pouch users prompted us to examine whether the association between using pouches as a cessation aid and quit success differed according to whether participants also used e-cigarettes as a cessation aid during their quit attempt. There was an interaction, with pouch use associated with higher quit success among those who did not use e-cigarettes during their quit attempt, but no clear evidence of an association among those who did. This could indicate that nicotine pouches do not provide substantial additional benefit when used alongside e-cigarettes, which are among the most effective smoking cessation aids,^14,15^ but may be associated with greater odds of quit success when used in the absence of an established alternative cessation aid. For people concerned about the potential harms of e-cigarettes (many people who smoke perceive them to be as harmful as cigarettes^27^), nicotine pouches may be a more appealing alternative because they deliver nicotine without inhalation into the lungs.

Taken together, our findings provide uncertain evidence about the association between nicotine pouch use as a cessation aid and smoking cessation. The adjusted estimate was consistent with a modest benefit (OR=1.15), although the confidence interval included the possibility of no effect, a modest adverse effect, or a larger benefit (95% CI 0.80–1.65), while excluding very large effects in either direction. There was, however, evidence of a benefit among those who did not use e-cigarettes in their quit attempt, with the estimate (OR=1.91, 95% CI 1.23–2.97) potentially similar to that observed for e-cigarettes in randomised controlled trials^15^ or real-world observational studies^7,28^, although also compatible with the smaller effects generally observed for NRT.^7,14,29^

Our findings add to a limited existing evidence base: the Cochrane review identified only four small randomised trials (total *n*=284), with low or very low certainty evidence on smoking cessation outcomes.^9^ Further well-designed randomised controlled trials are therefore needed to determine whether, in what circumstances, and for whom, nicotine pouches improve smoking cessation outcomes, how they compare with other established cessation aids, and whether they provide additional benefit when used alongside them.

This study has several strengths, including the large, nationally representative sample and monthly data collection over more than five years. However, several limitations should be considered. First, residual confounding is possible despite adjustment for a broad range of sociodemographic and smoking-related characteristics. Second, although we adjusted for concurrent use of other cessation aids, we did not capture their frequency, duration, or timing within the quit attempt. This limited our ability to determine how pouches were used alongside other products. Third, strength of urges to smoke was measured at the time of survey rather than at the start of the quit attempt and may therefore have been affected by subsequent smoking and quit success. When strength of urges to smoke was excluded from the adjusted model, the association between pouch use and quit success was larger but remained imprecisely estimated. Finally, the number of pouch users was relatively small, particularly for some subgroup analyses, resulting in imprecise estimates. As pouch use continues to increase, larger numbers of users will allow more precise estimates of effectiveness and assessment of whether effectiveness varies across different subgroups.

In conclusion, use of nicotine pouches as a smoking cessation aid has increased rapidly in Great Britain, particularly among younger adults and men. Pouches were commonly used alongside other cessation aids. Although the overall association between pouch use and quit success was uncertain after adjustment, pouch use was associated with higher quit success among those who did not also use e-cigarettes during their quit attempt, suggesting a potential role for pouches among people not already using an established cessation aid. As use continues to grow, larger observational studies will provide more precise estimates across population groups, while randomised trials are needed to determine whether nicotine pouches improve cessation outcomes and whether they offer benefits beyond established cessation aids.

## Data Availability

All data produced in the present study are available upon reasonable request to the authors.

## Declarations

### Ethics approval

Ethical approval for the STS was granted originally by the UCL Ethics Committee (ID 0498/001). Participants provide informed consent to take part in the study, and all methods are carried out in accordance with relevant regulations. The data are not collected by UCL and are anonymised when received by UCL.

