## Supplementary material for "Nicotine pouch use for smoking cessation in Great Britain: real-world effectiveness, trends, and user characteristics, 2021–2026": Table S1

**Table S1.** Model selection

| | 3 knots, AIC | 4 knots, AIC | ΔAIC (4 vs. 3 knots) | Selected model |
|---|---|---|---|---|
| All adults who attempted to quit smoking in the past year | 2562.09 | 2559.56 | -2.53 | 4 knots |
| By age | 2448.88 | 2449.68 | +0.80 | 3 knots |
| By gender | 2464.39 | 2463.56 | -0.83 | 3 knots |
| By occupational social grade | 2568.86 | 2566.15 | -2.71 | 4 knots |

AIC, Akaike Information Criterion. ΔAIC is calculated as AIC for the 4-knot model minus AIC for the 3-knot model.
The best fitting model was selected as the model with the lowest AIC or the simplest model within 2 AIC units.

**Table S2.** Sample demographic characteristics and strength of urges to smoke

| | **Column %**[1] |
|---|---|
| *Unweighted N* | 7,995 |
| Age (years) | |
| Mean (SD) | 39.0 (15.7) |
| 18-24 | 22.1 |
| 25-34 | 28.0 |
| 35-44 | 17.2 |
| 45-54 | 13.2 |
| 55-64 | 11.4 |
| ≥65 | 8.1 |
| *Missing, n* | 0 |
| Gender | |
| Men | 52.0 |
| Women | 46.8 |
| Other | 1.2 |
| *Missing, n* | 33 |
| Socioeconomic position | |
| ABC1 (more advantaged) | 42.1 |
| C2DE (less advantaged) | 57.9 |
| *Missing, n* | 0 |
| Strength of urges to smoke | |
| Mean (SD) | 1.6 (1.3) |
| 0. Not at all | 28.1 |
| 1. Slight | 18.6 |
| 2. Moderate | 31.6 |
| 3. Strong | 13.9 |
| 4. Very strong | 5.0 |
| 5. Extremely strong | 2.9 |
| *Missing, n* | 147 |

[1] Data are presented as weighted percentages, unless otherwise indicated. Sample sizes and numbers of missing cases are reported unweighted.

**Table S3.** Quitting characteristics

| | **Column %[1]** |
|---|---|
| *Unweighted N* | 7,995 |
| Successfully stopped smoking | 27.3 |
| *Missing, n* | 0 |
| Quit attempt was… | |
| Abrupt | 52.1 |
| Gradual | 47.9 |
| *Missing, n*[3] | 150 |
| Planned | 40.1 |
| Unplanned | 59.9 |
| *Missing, n*[3] | 341 |
| Quit attempt involved the use of … | |
| Nicotine pouches | 3.9 |
| E-cigarettes | 36.6 |
| NRT bought over-the-counter | 17.6 |
| Prescription pharmacotherapy[2] | 7.5 |
| Face-to-face behavioural support | 3.7 |
| *Missing, n* | 0 |
| Time since most recent quit attempt began (months) | |
| ≤1 | 15.1 |
| >1 and ≤3 | 22.2 |
| >3 and ≤6 | 23.9 |
| >6 and ≤12 | 38.9 |
| *Missing, n*[3] | 144 |
| Number of past-year quit attempts | |
| 1 | 61.3 |
| 2 | 20.5 |
| 3 | 9.9 |
| ≥4 | 8.3 |
| *Missing, n* | 0 |

[1] Data are presented as weighted percentages, unless otherwise indicated. Sample sizes and numbers of missing cases are reported unweighted.
[2] NRT, varenicline, bupropion, or cytisine.
[3] All missing cases on this variable were participants who responded that they did not know.

**Table S4.** Modelled estimates of the change in prevalence of nicotine pouch use as a smoking cessation aid among adults in Great Britain who made a past-year smoking quit attempt from June 2021 to July 2026: by age and gender

| | Prevalence, % [95% CI][1] | |
|---|---|---|
| | **June 2021** | **July 2026** |
| Men by age (years)[2] | | |
| 18-24 | 3.1 [1.0–8.7] | 26.1 [17.9–36.5] |
| 25-34 | 1.9 [0.7–4.6] | 7.8 [4.8–12.5] |
| 35-44 | 1.2 [0.3–4.7] | 3.4 [1.8–6.4] |
| 45-54 | 0.8 [0.2–4.2] | 2.8 [1.5–5.1] |
| 55-64 | 0.6 [0.1–4.9] | 3.4 [1.6–7.1] |
| ≥65 | 0.4 [0.0–13.6] | 6.2 [1.3–25.3] |
| Women by age (years)[2] | | |
| 18-24 | 1.2 [0.3–4.3] | 7.9 [2.9–19.7] |
| 25-34 | 1.8 [0.8–4.2] | 6.0 [2.9–11.9] |
| 35-44 | 2.2 [0.8–5.5] | 3.9 [1.8–8.2] |
| 45-54 | 1.9 [0.8–4.6] | 2.3 [1.1–4.9] |
| 55-64 | 1.5 [0.5–4.1] | 1.3 [0.4–4.2] |
| ≥65 | 0.8 [0.1–7.5] | 0.4 [0.0–6.2] |

CI, confidence interval. PR, prevalence ratio. Ref, reference category.
[1] Model-predicted prevalence for June 2021 and July 2026 from logistic regression models including a three-way interaction between age, gender, and time (survey month), with time modelled non-linearly using restricted cubic splines (three knots). Full trends are shown in **Figure S1**.
[2] Age modelled continuously using restricted cubic splines using data from people of all ages, with an interaction between age and time included. Estimates for each age group were taken from the mean exact age of individuals in that age range.

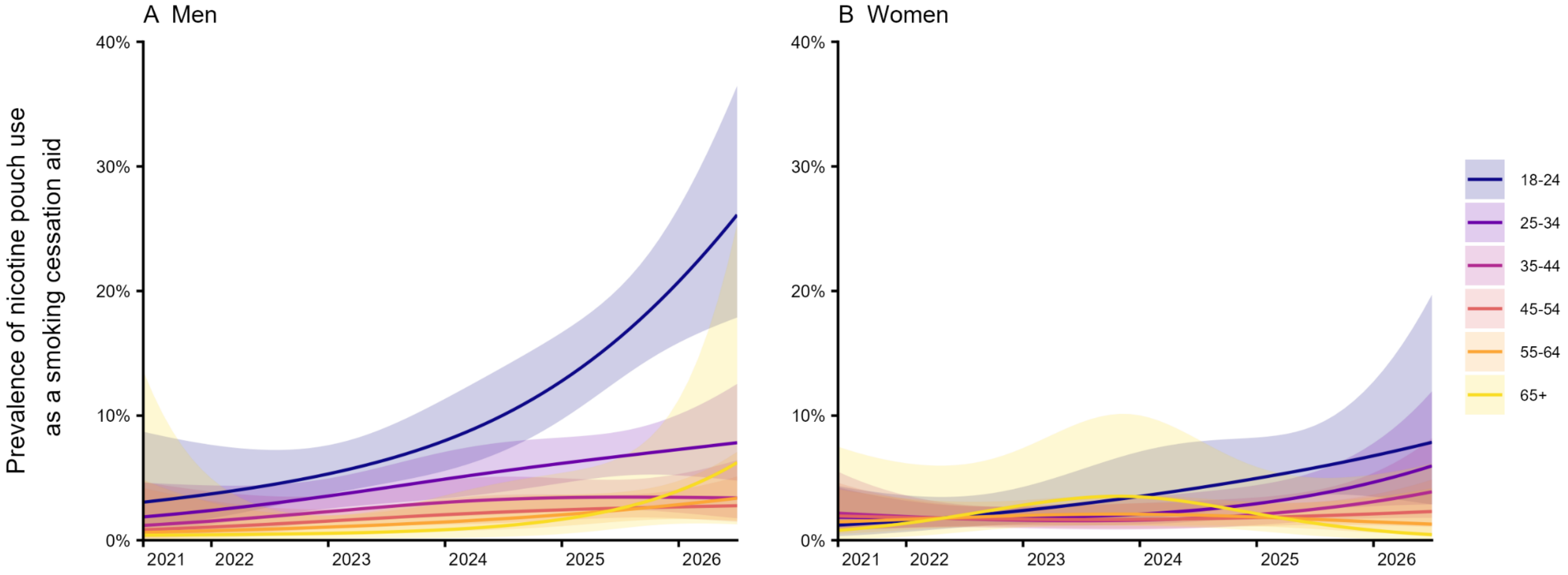


**Figure S1. Trends in the prevalence of nicotine pouch use as a smoking cessation aid by age and gender in Great Britain.**
Lines represent the modelled weighted prevalence by monthly survey wave (estimated using logistic regression, with time modelled with restricted cubic splines; three knots), shaded bands represent 95% CIs, and points represent the unmodelled weighted prevalence in each month. See **Table S4** for exact estimates in the first and last months of this period.
